# Health Condition and Test Availability as Predictors of Adults’ Mental Health during the COVID-19 Pandemic

**DOI:** 10.1101/2020.06.21.20137000

**Authors:** Huiyang Dai, Stephen X. Zhang, Kim Hoe Looi, Rui Su, Jizhen Li

**Author notes:** Corresponding author: Stephen X. Zhang; Address: 9-28 Nexus10 Tower, 10 Pulteney St, Adelaide SA 5000, Australia.

## Abstract

**Background:** Research identifying adults’ mental health during the COVID-19 pandemic relies solely on demographic predictors without examining adults’ health status during the COVID-19 pandemic as a potential predictor.

**Methods:** An online survey of 669 adults in Malaysia was conducted during May 2–8, 2020, six weeks after a Movement Control Order (MCO) was issued.

**Findings:** Adults’ health condition had curvilinear relationships (horizontally reversed J-shaped) with insomnia, anxiety, depression and distress. Reported test availability for COVID-19 (from “strongly disagree” to “strongly agree”) also had curvilinear relationships (horizontally reversed J-shaped) with anxiety and depression. Younger adults reported worse mental health, but people from various religions and ethnic groups did not differ significantly in reported mental health.

**Interpretation:** Adults with worse health conditions had more mental health problems, especially adults at the lower end of the health spectrum. Test availability negatively predicted anxiety and depression, especially for adults experiencing poor COVID-19 test availability. The significant predictions of health condition and COVID-19 test availability suggest a new direction for the literature to identify psychiatric risk factors directly from health related variables during a pandemic.

**Funding:** Tsinghua University-INDITEX Sustainable Development Fund (Project No. TISD201904).

## 1. Introduction

In May 2020, the UN Secretary-General issued a message that the COVID-19 pandemic had resulted in massive mental suffering and called for actions.^1^ A number of studies have predicted the mental health of adults by means of demographic variables,^2–5^ but little research has predicted mental health based on adults’ health status during the COVID-19 pandemic. This study is the first, to the best of our knowledge, to identify such predictors, specifically adults’ health condition and availability of testing for COVID-19.

First, good health condition can lower individuals’ chance of COVID-19 infection, and healthcare workers (e.g., general practitioners) may already have some knowledge of the health status of people under their care. Second, as individuals have heterogeneous access to COVID-19 testing due to limited testing capacity in many countries, individuals who have poorer access to COVID-19 testing may be more concerned or anxious about the COVID-19 pandemic. Test availability for COVID-19 is potentially a unique predictor of mental health during the COVID-19 pandemic.

## 2. Methods

The first case of COVID-19 in Malaysia was confirmed on February 4, 2020, and on March 18, 2020, Malaysia implemented a Movement Control Order (MCO) to ban citizens from non-essential travel and mass gatherings. The data for this study was collected by an online survey from May 2 to 8, 2020, six weeks after the implementation of the MCO. On May 8, 2020, there were a total of 6,535 confirmed cases of COVID-19 and 108 deaths.^6^

Even though we were not aiming for a fully representative sample, we applied two-stage stratified sampling in terms of ethnicity, gender, age, and geographical area^7^ to ensure coverage of all regions and ethnic groups in Malaysia.^8^ Participation in this survey was voluntary and participants could opt-out at any time. Moreover, participants were assured anonymity and confidentiality of their responses. The survey was granted ethical approval by Tsinghua University (20200322). The online survey was issued in Malay, Mandarin and English, the major languages used in Malaysia.

The participants reported their demographic characteristics, including gender, age, education level, number of children under 18 years old in the household, religion, and ethnic group. We assessed health condition using the global health measure SF-1^9^ with a five-point scale from 1 to 5 (poor, fair, good, very good, excellent). To capture availability of testing for COVID-19 in Malaysia, we asked participants to rate the statement “I can get a test for COVID-19 rapidly if I need it” from 1 to 7 (strongly disagree, disagree, somewhat disagree, neither agree nor disagree, somewhat agree, agree, strongly agree). We used four dimensions for mental health: insomnia, anxiety, depression and distress.

### Insomnia

Adults’ insomnia was measured with the five-item Athens Insomnia Scale (AIS-5)^10^, including “I have trouble falling asleep” and “I feel tired and worn-out after my usual amount of sleep”. The items were scored from 1 (to a very small extent) to 5 (to a very large extent). The Cronbach’s alpha was 0·82.

### Anxiety

Adults’ anxiety was measured by the seven-item generalized anxiety disorder (GAD-7)^11^ scale. The seven items were scored from 0 (not at all) to 3 (nearly every day). The Cronbach’s alpha was 0·92.

### Depression

Adults’ depression was measured with the nine-item Patient Health Questionnaire depression module (PHQ-9),^12^ with items scored from 0 (not at all) to 3 (nearly every day). The Cronbach’s alpha was 0.90.

### Distress

Adults’ psychological distress was measured with the six-item K6 screening scale,^13^ with items scored from 1 (all of the time) to 5 (none of the time). The Cronbach’s alpha was 0·95.

We applied ordinary least squares (OLS) regression for the four dependent variables. Stata 16·0 was used to identify the predictors for insomnia, anxiety, depression and distress.

## 3. Results

### 3.1 Descriptive findings

Overall, 669 adults from all the states and federal territories of Malaysia participated in this survey. Table 1 shows the descriptive statistics of the sample. Participation by both genders was almost equal. The youngest participant was 21 years old and the oldest was 71 years old. Malaysia is a diverse country in terms of ethnicity and religion. Religion and ethnicity were reported in Table 1. Overall, our sample captured all the major ethnic and religious groups in Malaysia, but the sample is not taken as representative.

**Table 1.**
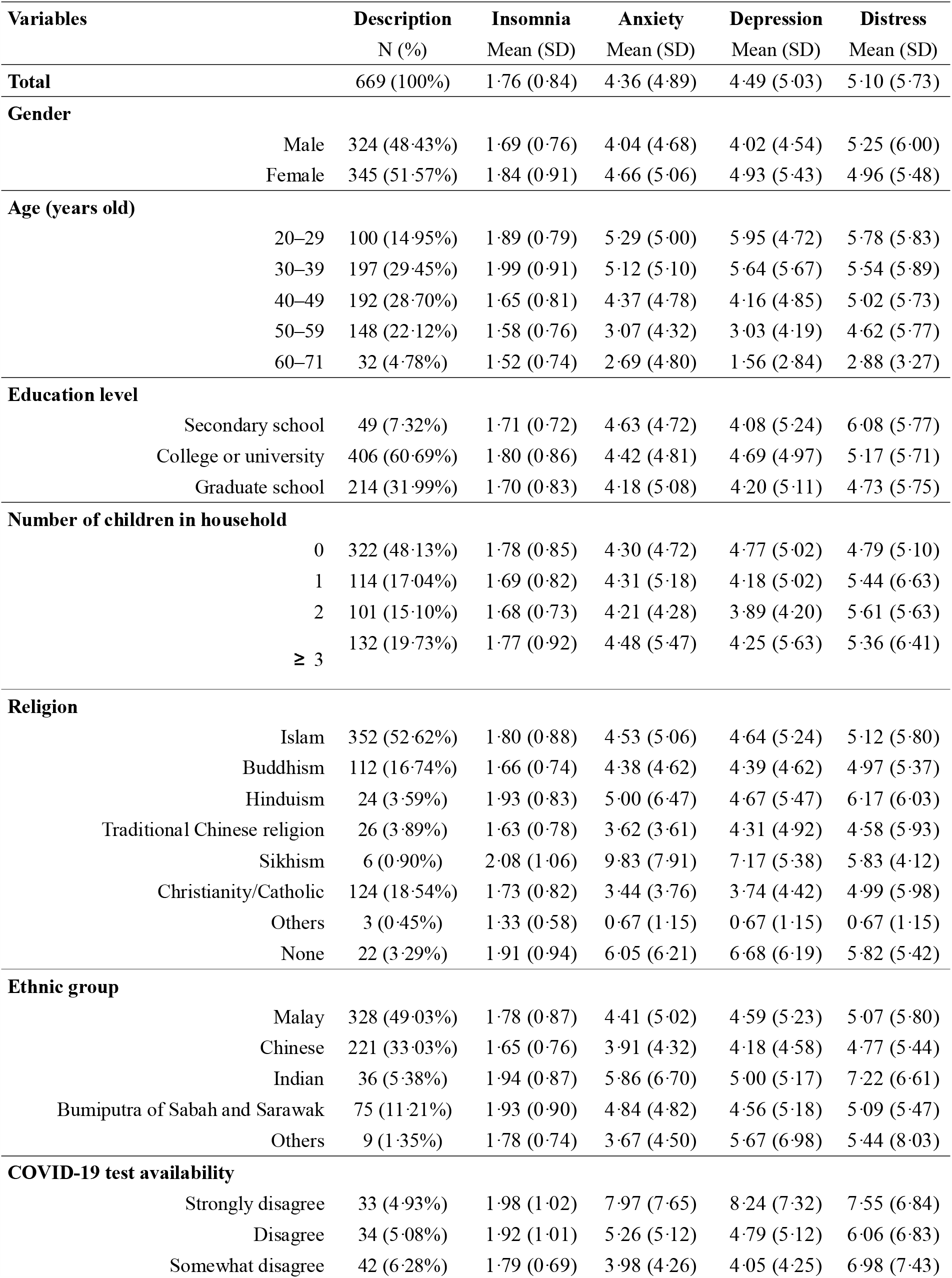
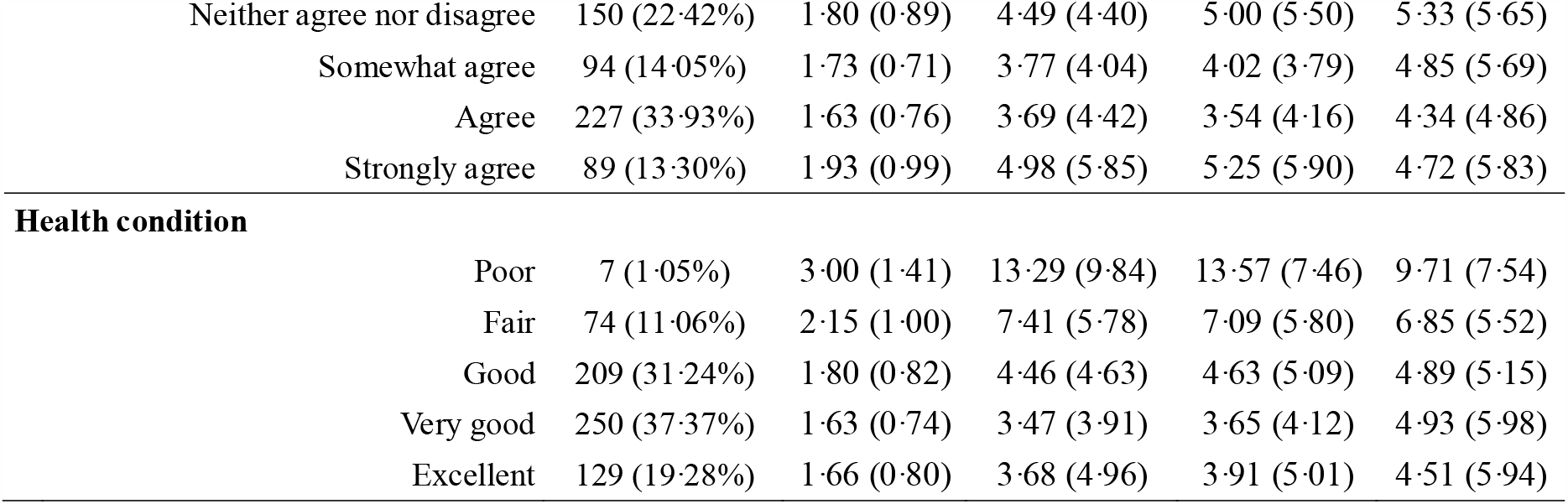
Descriptive statistics of Malaysian participants

| Variables |  | Description<br>N (%) | Insomnia<br>Mean (SD) | Anxiety<br>Mean (SD) | Depression<br>Mean (SD) | Distress<br>Mean (SD) |
| --- | --- | --- | --- | --- | --- | --- |
| **Total** |  | 669 (100%) | 1.76 (0.84) | 4.36 (4.89) | 4.49 (5.03) | 5.10 (5.73) |
| **Gender** |  |  |  |  |  |  |
|  | Male | 324 (48.43%) | 1.69 (0.76) | 4.04 (4.68) | 4.02 (4.54) | 5.25 (6.00) |
|  | Female | 345 (51.57%) | 1.84 (0.91) | 4.66 (5.06) | 4.93 (5.43) | 4.96 (5.48) |
| **Age (years old)** |  |  |  |  |  |  |
|  | 20–29 | 100 (14.95%) | 1.89 (0.79) | 5.29 (5.00) | 5.95 (4.72) | 5.78 (5.83) |
|  | 30–39 | 197 (29.45%) | 1.99 (0.91) | 5.12 (5.10) | 5.64 (5.67) | 5.54 (5.89) |
|  | 40–49 | 192 (28.70%) | 1.65 (0.81) | 4.37 (4.78) | 4.16 (4.85) | 5.02 (5.73) |
|  | 50–59 | 148 (22.12%) | 1.58 (0.76) | 3.07 (4.32) | 3.03 (4.19) | 4.62 (5.77) |
|  | 60–71 | 32 (4.78%) | 1.52 (0.74) | 2.69 (4.80) | 1.56 (2.84) | 2.88 (3.27) |
| **Education level** |  |  |  |  |  |  |
|  | Secondary school | 49 (7.32%) | 1.71 (0.72) | 4.63 (4.72) | 4.08 (5.24) | 6.08 (5.77) |
|  | College or university | 406 (60.69%) | 1.80 (0.86) | 4.42 (4.81) | 4.69 (4.97) | 5.17 (5.71) |
|  | Graduate school | 214 (31.99%) | 1.70 (0.83) | 4.18 (5.08) | 4.20 (5.11) | 4.73 (5.75) |
| **Number of children in household** |  |  |  |  |  |  |
|  | 0 | 322 (48.13%) | 1.78 (0.85) | 4.30 (4.72) | 4.77 (5.02) | 4.79 (5.10) |
|  | 1 | 114 (17.04%) | 1.69 (0.82) | 4.31 (5.18) | 4.18 (5.02) | 5.44 (6.63) |
|  | 2 | 101 (15.10%) | 1.68 (0.73) | 4.21 (4.28) | 3.89 (4.20) | 5.61 (5.63) |
|  | ≥ 3 | 132 (19.73%) | 1.77 (0.92) | 4.48 (5.47) | 4.25 (5.63) | 5.36 (6.41) |
| **Religion** |  |  |  |  |  |  |
|  | Islam | 352 (52.62%) | 1.80 (0.88) | 4.53 (5.06) | 4.64 (5.24) | 5.12 (5.80) |
|  | Buddhism | 112 (16.74%) | 1.66 (0.74) | 4.38 (4.62) | 4.39 (4.62) | 4.97 (5.37) |
|  | Hinduism | 24 (3.59%) | 1.93 (0.83) | 5.00 (6.47) | 4.67 (5.47) | 6.17 (6.03) |
|  | Traditional Chinese religion | 26 (3.89%) | 1.63 (0.78) | 3.62 (3.61) | 4.31 (4.92) | 4.58 (5.93) |
|  | Sikhism | 6 (0.90%) | 2.08 (1.06) | 9.83 (7.91) | 7.17 (5.38) | 5.83 (4.12) |
|  | Christianity/Catholic | 124 (18.54%) | 1.73 (0.82) | 3.44 (3.76) | 3.74 (4.42) | 4.99 (5.98) |
|  | Others | 3 (0.45%) | 1.33 (0.58) | 0.67 (1.15) | 0.67 (1.15) | 0.67 (1.15) |
|  | None | 22 (3.29%) | 1.91 (0.94) | 6.05 (6.21) | 6.68 (6.19) | 5.82 (5.42) |
| **Ethnic group** |  |  |  |  |  |  |
|  | Malay | 328 (49.03%) | 1.78 (0.87) | 4.41 (5.02) | 4.59 (5.23) | 5.07 (5.80) |
|  | Chinese | 221 (33.03%) | 1.65 (0.76) | 3.91 (4.32) | 4.18 (4.58) | 4.77 (5.44) |
|  | Indian | 36 (5.38%) | 1.94 (0.87) | 5.86 (6.70) | 5.00 (5.17) | 7.22 (6.61) |
|  | Bumiputra of Sabah and Sarawak | 75 (11.21%) | 1.93 (0.90) | 4.84 (4.82) | 4.56 (5.18) | 5.09 (5.47) |
|  | Others | 9 (1.35%) | 1.78 (0.74) | 3.67 (4.50) | 5.67 (6.98) | 5.44 (8.03) |
| **COVID-19 test availability** |  |  |  |  |  |  |
|  | Strongly disagree | 33 (4.93%) | 1.98 (1.02) | 7.97 (7.65) | 8.24 (7.32) | 7.55 (6.84) |
|  | Disagree | 34 (5.08%) | 1.92 (1.01) | 5.26 (5.12) | 4.79 (5.12) | 6.06 (6.83) |
|  | Somewhat disagree | 42 (6.28%) | 1.79 (0.69) | 3.98 (4.26) | 4.05 (4.25) | 6.98 (7.43) |

|  |  |  |  |  |  |
| --- | --- | --- | --- | --- | --- |
| Neither agree nor disagree | 150 (22.42%) | 1.80 (0.89) | 4.49 (4.40) | 5.00 (5.50) | 5.33 (5.65) |
| Somewhat agree | 94 (14.05%) | 1.73 (0.71) | 3.77 (4.04) | 4.02 (3.79) | 4.85 (5.69) |
| Agree | 227 (33.93%) | 1.63 (0.76) | 3.69 (4.42) | 3.54 (4.16) | 4.34 (4.86) |
| Strongly agree | 89 (13.30%) | 1.93 (0.99) | 4.98 (5.85) | 5.25 (5.90) | 4.72 (5.83) |
| **Health condition** |  |  |  |  |  |
| Poor | 7 (1.05%) | 3.00 (1.41) | 13.29 (9.84) | 13.57 (7.46) | 9.71 (7.54) |
| Fair | 74 (11.06%) | 2.15 (1.00) | 7.41 (5.78) | 7.09 (5.80) | 6.85 (5.52) |
| Good | 209 (31.24%) | 1.80 (0.82) | 4.46 (4.63) | 4.63 (5.09) | 4.89 (5.15) |
| Very good | 250 (37.37%) | 1.63 (0.74) | 3.47 (3.91) | 3.65 (4.12) | 4.93 (5.98) |
| Excellent | 129 (19.28%) | 1.66 (0.80) | 3.68 (4.96) | 3.91 (5.01) | 4.51 (5.94) |

The average levels of mental health of adults in Malaysia were different from adults in other countries. The mean scores of insomnia (AIS-5), anxiety (GAD-7), depression (PHQ-9) and distress (K6) were 1·76 (SD = 0·84), 4·50 (SD = 4·90), 4·67 (SD = 5·12), and 5·95 (SD = 6·51), respectively. The mean scores of depression and anxiety in Malaysia were significantly lower than those in a sample of 300 adults collected on January 31 to February 7, 2020 in China^14^ of 8·3 (difference = -3·63, se = 0·39, 95% CI: -2·95 to -4·45, *p* < 0·001) and 7·7 (difference = -3·20, se = 0·37, 95% CI: -2·47 to -3·93, *p* < 0·001), and also lower than scores in a sample of 1,009 adults on April 10–20, 2020 in Austria^15^ of 6·20 (difference = -1·53, se = 0·26, 95% CI: -0·98 to -2·02, *p* < 0·001) and 5·85 (difference = -1·35, se = 0·26, 95% CI: -0·98 to -2·02, *p* < 0·001). The average degree of distress in a sample of 369 working adults (mean = 8·46) in China^16^ was significantly higher than in our sample (difference = 3·36, se = 0·32, 95% CI: 3·98 to 2·74, *p* < 0·001). The proportion of insomnia disorder in our sample (38·9%, n = 669) was similar to that in a sample collected during April 10–13, 2020 in Greece (37·6%, n = 2,427, difference = 1·3%, *p* = 0·540) during the COVID-19 pandemic.^17^

### 3.2 Predictors of insomnia, anxiety, depression, and distress

Table 2 presents the results of the regression models. The quadratic terms of health condition (health condition – square in Table 2) were significant across the regressions for all four dimensions of mental health: insomnia (β = 0·10, 95% CI: 0·03 to 0·17, *p* = 0·003), anxiety (β = 0·79, 95% CI: 0·36 to 1·22, *p* < 0·001), depression (β = 0·74, 95% CI: 0·38 to 1·11, *p* < 0·001), and distress (β = 0·45, 95% CI: 0·01 to 0·88, *p* = 0·045), demonstrating curvilinear relationships between health condition and mental health dimensions. The margin analysis of the slope of insomnia by health condition was -0·71 (*p* < 0·001) at “poor”, -0·51 (*p* < 0·001) at “fair”, -0·31 (*p* < 0·001) at “good”, -0·10 (*p* = 0·017) at “very good”, and 0·10 (*p* = 0·328) at “excellent” health condition, showing a horizontally reversed J-shaped curve across the scoring range of health condition· Similar curvilinear relationships were also observed from “poor” to “excellent” health condition for anxiety (d = -5·34, *p* < 0·001; d = -3·76, *p* < 0·001; d = -2·18, *p* < 0·001; d = -0·60, *p* = 0·019; d = 0·98, *p* = 0·114), depression (d = -5·04, *p* < 0·001; d = -3·55, *p* < 0·001; d = -2·06, *p* < 0·001; d = -0·57, *p* = 0·024; d = 0·91, *p* = 0·104) and distress (d = -3·11, *p* = 0·006; d = -2·21, *p* = 0·002; d = -1·32, *p* < 0·001; d = -0·43, *p* = 0·188; d = 0·46, *p* = 0·511).

**Table 2.** Predictors of adults’ insomnia, anxiety, depression and distress by regression analyses (N = 669)

| Variables | Insomnia |  | Anxiety |  | Depression |  | Distress |  |
| --- | --- | --- | --- | --- | --- | --- | --- | --- |
| | $\beta$ (95% CI) | *p*-value | $\beta$ (95% CI) | *p*-value | $\beta$ (95% CI) | *p*-value | $\beta$ (95% CI) | *p*-value |
| **Health condition–square** | 0.10 (0.03 to 0.17) | 0.003 | 0.79 (0.36 to 1.22) | 0.000 | 0.74 (0.38 to 1.11) | 0.000 | 0.45 (0.01 to 0.88) | 0.045 |
| **Health condition** | -0.91 (-1.41 to -0.42) | 0.000 | -6.91 (-10.06 to -3.77) | 0.000 | -6.53 (-9.18 to -3.88) | 0.000 | -4.00 (-7.08 to -0.91) | 0.011 |
| **Test availability–square** | 0.01 (-0.01 to 0.03) | 0.404 | 0.17 (0.03 to 0.31) | 0.017 | 0.16 (0.02 to 0.29) | 0.027 | 0.02 (-0.14 to 0.18) | 0.790 |
| **Test availability** | -0.11 (-0.30 to 0.08) | 0.253 | -1.82 (-3.10 to -0.55) | 0.005 | -1.73 (-2.99 to -0.46) | 0.007 | -0.67 (-2.08 to 0.75) | 0.355 |
| **Gender (Female)** | 0.08 (-0.05 to 0.21) | 0.214 | 0.22 (-0.48 to 0.93) | 0.535 | 0.49 (-0.24 to 1.22) | 0.190 | -0.54 (-1.40 to 0.32) | 0.217 |
| **Age** | -0.02 (-0.02 to -0.01) | 0.000 | -0.08 (-0.11 to -0.05) | 0.000 | -0.11 (-0.14 to -0.08) | 0.000 | -0.06 (-0.10 to -0.02) | 0.005 |
| **Education level** | -0.05 (0.36 to -0.15) | 0.056 | -0.25 (-0.86 to 0.36) | 0.414 | -0.10 (-0.74 to 0.55) | 0.770 | -0.64 (-1.40 to 0.12) | 0.100 |
| **Number of children** | 0.02 (-0.02 to 0.07) | 0.331 | 0.16 (-0.09 to 0.41) | 0.209 | 0.02 (-0.27 to 0.30) | 0.912 | 0.25 (-0.07 to 0.56) | 0.131 |
| **Religion** |  |  |  |  |  |  |  |  |
| Islam | Reference group |  | Reference group |  | Reference group |  | Reference group |  |
| Buddhism | -0.12 (-0.54 to 0.31) | 0.593 | -0.02 (-2.39 to 2.35) | 0.986 | 0.61 (-1.75 to 2.96) | 0.613 | 0.15 (-2.66 to 2.97) | 0.916 |
| Hinduism | 0.08 (-0.62 to 0.79) | 0.814 | -0.02 (-4.10 to 4.07) | 0.994 | 1.13 (-2.77 to 5.04) | 0.569 | -3.34 (-10.00 to 3.32) | 0.325 |
| Traditional Chinese religion | -0.15 (-0.64 to 0.35) | 0.561 | -0.80 (-3.29 to 1.69) | 0.529 | 0.71 (-2.09 to 3.51) | 0.618 | -0.37 (-3.89 to 3.16) | 0.837 |
| Sikhism | 0.11 (-0.8 to 1.03) | 0.807 | 3.78 (-2.57 to 10.13) | 0.243 | 3.31 (-1.23 to 7.84) | 0.153 | -4.90 (-11.86 to 2.07) | 0.168 |
| Christianity/Catholic | -0.12 (-0.51 to 0.27) | 0.546 | -1.56 (-3.71 to 0.58) | 0.154 | -0.22 (-2.39 to 1.95) | 0.840 | -0.16 (-2.65 to 2.33) | 0.900 |
| Others | -0.77 (-1.89 to 0.34) | 0.175 | -6.10 (-11.52 to -0.69) | 0.027 | -5.64 (-10.54 to -0.74) | 0.024 | -6.22 (-10.94 to -1.50) | 0.010 |
| None | 0.12 (-0.40 to 0.64) | 0.642 | 1.40 (-1.90 to 4.69) | 0.406 | 2.70 (-0.46 to 5.85) | 0.094 | 0.42 (-2.98 to 3.83) | 0.807 |
| **Ethnic group** |  |  |  |  |  |  |  |  |
| Malay | Reference group |  | Reference group |  | Reference group |  | Reference group |  |
| Chinese | -0.03 (-0.45 to 0.39) | 0.873 | -0.03 (-2.30 to 2.23) | 0.977 | -1.08 (-3.33 to 1.16) | 0.343 | -0.31 (-2.98 to 2.37) | 0.823 |
| Indian | 0.17 (-0.49 to 0.82) | 0.622 | 1.24 (-2.30 to 4.78) | 0.491 | -0.78 (-4.17 to 2.61) | 0.651 | 5.30 (-1.23 to 11.84) | 0.111 |
| Bumiputra of Sabah and Sarawak | 0.19 (-0.19 to 0.58) | 0.329 | 1.25 (-0.91 to 3.41) | 0.256 | -0.38 (-2.52 to 1.77) | 0.731 | 0.07 (-2.22 to 2.36) | 0.951 |
| Others | 0.24 (-0.32 to 0.80) | 0.405 | 0.80 (-2.59 to 4.20) | 0.642 | 1.59 (-2.34 to 5.52) | 0.427 | 2.16 (-4.09 to 8.42) | 0.497 |

The quadratic term of test availability (test availability – square in Table 2) was positively associated with anxiety (β = 0·17, 95% CI: 0·03 to 0·31, *p* = 0·017) and depression (β = 0·16, 95% CI: 0·02 to 0·29, *p* = 0·027). The margin analysis of the slope of anxiety by test availability was -1·49 (*p* < 0·004) at “strongly disagree”, -1·15 (*p* = 0·002) at “disagree”, -0·81 (*p* = 0·001) at “somewhat disagree”, -0·48 (*p* = 0·001) at “neither disagree nor agree”, -0·14 (*p* = 0·311) at “somewhat agree”, 0·19 (*p* = 0·423) at “agree”, and 0·53 (*p* = 0·152) at “strongly agree”, showing a horizontally reversed J-shaped curve across the scoring range of test availability. The margin analysis of the slope of depression by test availability demonstrated a similar pattern at the 7-point anchor from “strongly disagree” to “strongly agree” (d = -1·41, *p* = 0·005; d = -1·10, *p* = 0·003; d = -0·79, *p* = 0·001; d = -0·48, *p* = 0·001; d = -0·17, *p* = 0·263; d = 0·16, *p* = 0·562; d = 0·46, *p* = 0·227).

In addition, age negatively predicted insomnia (β = -0·02, 95% CI: -0·02 to -0·01, *p* < 0·001), anxiety (β = -0·08, 95% CI: -0·11 to -0·05, *p* < 0·001), depression (β = -0·11, 95% CI: -0·14 to -0·08, *p* < 0·001) and distress (β = -0·06, 95% CI: -0·10 to -0·02, *p* = 0·005). Although we included religion and ethnicity in the regression model, insomnia, anxiety, depression and distress did not vary significantly by them· Significant results are shown in Figure 1.

**Figure 1.**
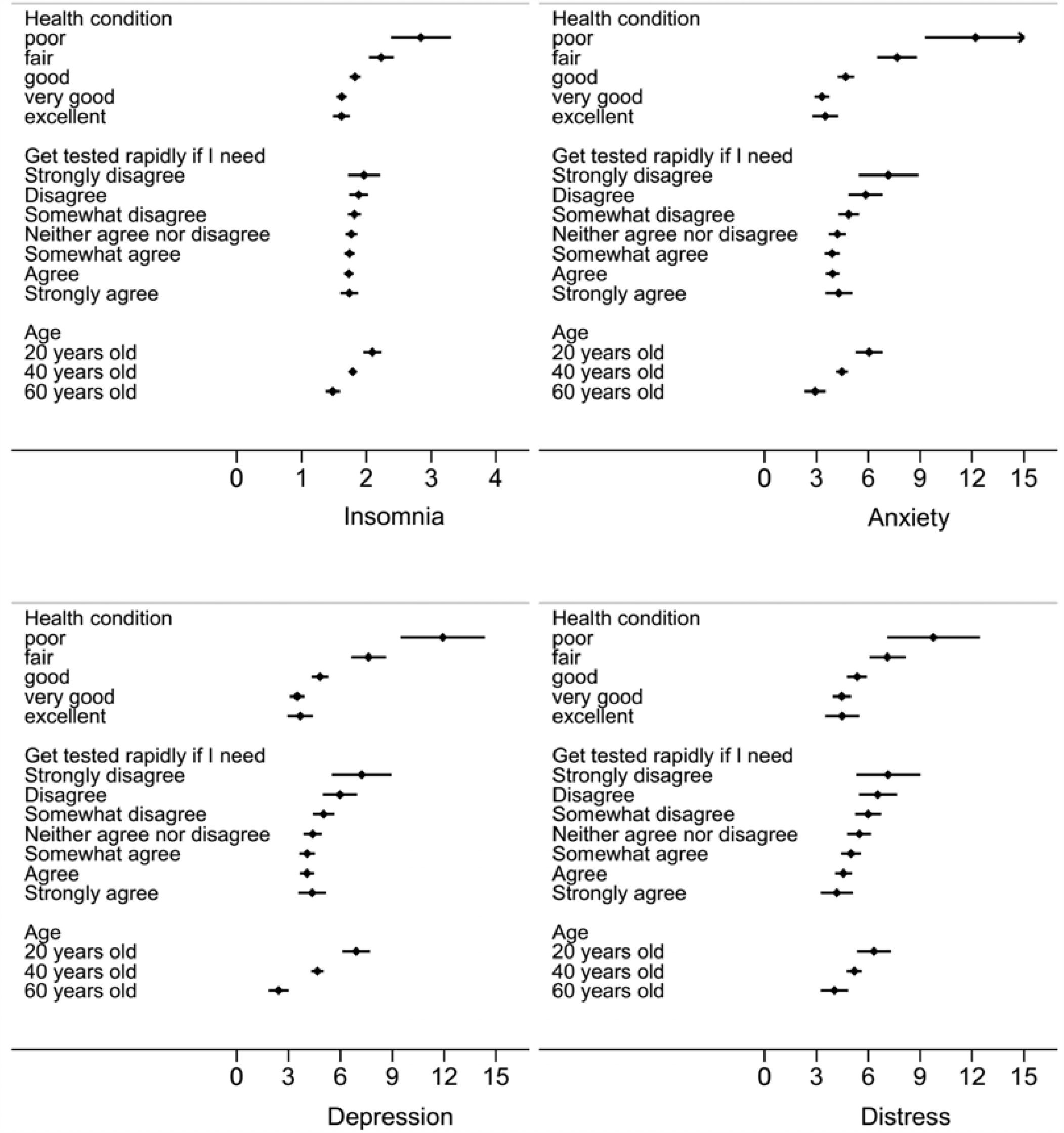
**Predicted value and 95% confidence intervals (CIs) of insomnia, anxiety, depression and distress by health condition, COVID-19 test availability and age**.

## 4. Discussion

This study identifies several predictors of insomnia, anxiety, depression, and distress among adults in Malaysia during the COVID-19 pandemic. Consistent with past studies,^5,18^ age was found to be a predictor of mental health problems for the general population in Malaysia. This is also consistent with studies of healthcare workers, where older healthcare workers were less likely to have mental health problems.^19,20^ However, other predictors found in the literature, such as education^16^ and gender,^17^ failed to predict mental health among adults in Malaysia, similar to a study in the UK.^5^ Our results suggest that future research should identify the effect of education and gender across more countries and future meta-analysis should identify contingent factors. Religion and ethnic group did not predict Malaysian adults’ mental health, consistent with another Malaysian study^21^ which found that ethnicity is not correlated with mental health disorder. In line with previous research, our findings highlight the need to identify specific predictors of mental health under various contexts of the COVID-19 pandemic.^19^

More importantly, this study uncovers two unique risk factors for mental health. The first risk factor is existing health condition, which had significant curvilinear relationships with insomnia, anxiety, depression and distress. The results indicated that adults with worse reported health condition had worse mental health, especially for those at the lower end of the health spectrum. In other words, individuals’ existing health condition is a useful screener of insomnia, anxiety, depression and distress for less healthy adults.

The second risk factor is test availability for COVID-19 having curvilinear relationships with anxiety and depression. Test unavailability predicted worse anxiety and depression, especially for people who disagreed that they could get tested for COVID-19 when needed. There was also no significant difference in mental health among people who “somewhat agreed”, “agreed” and “strongly agreed” that they could get a COVID-19 test. Test availability could be a predictor of mental health, especially for adults who reported they lack access to COVID-19 tests.

Our findings suggest that healthcare service providers could use adults’ general health condition and COVID-19 test availability to identify mentally vulnerable adults. The curvilinear relationships highlight the need to pay more attention to adults with poor health condition and adults who lack access to a COVID-19 test. Healthcare service providers such as hospitals may be able to use the health records of their patients and COVID-19 testing coverage to help identify those who need more mental health assistance.

### 4.1 Limitations and future work

Firstly, we used a two-stage stratified sampling method to aim for a broader coverage of Malaysian adults, but our study did not aim for a representative sample of adults in Malaysia during the COVID-19 pandemic. Secondly, we used SF-1, a brief one-item measure of general health condition, and future research may use the lengthier form of SF-12 or SF-36. Thirdly, instead of general health condition, future research may explore specific medical issues, such as heart disease, diabetes, or cancer, as predictors of mental health. Fourthly, we measured adults’ perceived test availability for COVID-19 because we were interested in their mental health and future research may use alternative indicators of COVID-19 test availability.

## 4.2 Conclusion

This study identified two unique predictors of health condition and test availability for COVID-19 for Malaysian adults’ mental health, operationalized as insomnia, anxiety, depression and distress. Unlike the demographic predictors identified in prior research, these two risk factors suggest new risk factors to predict mental health. Moreover, these predictors carried quadratic associations with various mental health dimensions, implying a need to focus on curvilinear predictors of mental health.

## Data Availability

The data for this study was collected by an online survey from May 2 to 8, 2020, six weeks after the implementation of the MCO.

## Contribution Statement

H. D.: Visualization, Writing – Original Draft, Writing – Review & Editing

S. X. Z.: Conceptualization, Investigation, Methodology, Formal analysis, Writing – Original, Writing - Review & Editing

K. H. L.: Investigation (data collection), Writing - Review & Editing

R. S.: Investigation (data collection)

J. L.: Resources, Funding acquisition

## ACKNOWLEDGEMENT

We acknowledge the support of Tsinghua University-INDITEX Sustainable Development Fund (Project No. TISD201904).

